# Estimating historical impacts of vaccination against influenza B/Yamagata in the United States to inform possible risks of re-emergence in the absence of vaccination

**DOI:** 10.1101/2024.10.30.24316093

**Authors:** Sinead E. Morris, Sarabeth M. Mathis, Jessie R. Chung, Brendan Flannery, Alissa O’Halloran, Catherine H. Bozio, Peng-Jun Lu, Tammy A. Santibanez, Peter Daly, Angiezel Merced-Morales, Krista Kniss, Alicia Budd, Lisa A. Grohskopf, Carrie Reed, Matthew Biggerstaff, A. Danielle Iuliano

**Affiliations:** Influenza Division, Centers for Disease Control and Prevention, Atlanta, GA, USA, 30329; Goldbelt Professional Services, Chesapeake, VA, USA, 23320; Immunization Services Division, Centers for Disease Control and Prevention, Atlanta, GA, USA, 30329

**Author notes:** **Disclaimer:** The findings and conclusions in this report are those of the authors and do not necessarily represent the views of the Centers for Disease Control and Prevention.

**Keywords:** vaccination, seasonal influenza, B/Yamagata, disease burden, vaccine-preventable burden

## Abstract

Influenza B/Yamagata viruses have not been detected globally since 2020 and will be removed from U.S. 2024/25 seasonal influenza vaccines. We inferred impacts of vaccination against B/Yamagata from 2016/17–2019/20 by combining B/Yamagata prevalence data with model-based estimates of disease burden prevented by vaccination against all influenza B viruses. B/Yamagata comprised approximately 16–22% of positive virus specimens in 2016/17 and 2017/18, compared to 1% in 2018/19 and 2019/20. Across all seasons, we estimated that vaccination against B/Yamagata prevented 4.15 million illnesses, 58,500 hospitalizations, and 4,070 deaths, and that 22.9 million B/Yamagata-associated illnesses, 340,000 hospitalizations, and 25,100 deaths would have occurred without vaccination. Vaccination prevented the most B/Yamagata hospitalizations among adults ≥65 years but prevented the greatest percentage of B/Yamagata hospitalizations among children 6 months–4 years. Our results may help assess the potential impact if B/Yamagata were to recirculate in the absence of vaccination.

## Introduction

Seasonal influenza vaccination is recommended for everyone ≥6 months who does not have contraindications in the United States (U.S.)^1^. Quadrivalent seasonal influenza vaccines which include antigens for two influenza B virus lineages, B/Yamagata and B/Victoria, have been available since 2013^2^. However, B/Yamagata viruses have not been detected globally since 2020 and will be removed from U.S. vaccines starting in the 2024/25 season (although global surveillance for B/Yamagata will continue)^3^. Understanding the historical impact of vaccination against B/Yamagata is important to help assess the potential public health consequences if B/Yamagata were to recirculate in the absence of vaccine-mediated protection.

The U.S. Centers for Disease Control and Prevention (CDC) estimates disease burden prevented by influenza vaccination each season using a compartmental framework that accounts for vaccine effectiveness (VE), vaccination coverage, and time-varying risks of influenza exposure^4^. We have demonstrated an extension of this framework that partitions burden prevented by vaccination between influenza A and influenza B viruses^5^, and hypothesized that the latter estimates could provide insights into the impact of vaccination against B/Yamagata when combined with information on levels of B/Yamagata circulation.

To explore this, we identified information on B/Yamagata circulation in the United States for four seasons preceding its disappearance (2016/17–2019/20)^6^. These seasons ranged in epidemic severity from moderate (2016/17, 2018/19 and 2019/20) to high (2017/18)^7^, and in levels of B/Yamagata circulation^8^. We then combined this information with our reported estimates of influenza-associated disease burden and burden prevented by vaccination for both influenza B lineages. We estimated age-specific numbers of illnesses, hospitalizations, and deaths prevented by vaccination against B/Yamagata, and the corresponding number that would have been caused by B/Yamagata without vaccination. Our results may help inform efforts to assess potential future landscapes of influenza disease burden in the context of changing influenza B epidemiology and absence of vaccine protection against B/Yamagata.

## Methods

### Surveillance data

We obtained information on the prevalence of influenza B/Yamagata from 2016/17–2019/20 from CDC’s FluView^6^. These data included the weekly number of influenza-positive specimens identified by virus type (influenza A or B), influenza A subtype, or influenza B lineage and reported by participating public health laboratories from the World Health Organization (WHO) Collaborating Laboratories System and National Respiratory and Enteric Virus Surveillance System (NREVSS). Data were available for five age groups: 0–4 years, 5–17 years, 18–49 years, 50–64 years, and ≥65 years. For each season, *s*, and age group, *a*, we calculated the prevalence of B/Yamagata among influenza B viruses, *p*_*a,s*_, as the proportion of all influenza B specimens with known lineage that were positive for B/Yamagata. We also calculated prevalence among all influenza viruses (i.e., including influenza A viruses in addition to influenza B viruses), as the proportion of all influenza specimens with known lineage or subtype that were positive for B/Yamagata.

### Framework for estimating burden prevented by vaccination for B/Yamagata

We previously estimated disease burden and burden prevented by vaccination for all influenza B lineages from 2016/17–2019/20 by extending a compartmental framework developed by the CDC^4,5^. These estimates were generated for four disease severity categories (illnesses, medically-attended illnesses, hospitalizations, and deaths) and five age groups (6 months–4 years, 5–17 years, 18–49 years, 50–64 years, and ≥65 years), and included uncertainty distributions obtained from Monte Carlo simulation.

For each age group (*a*) and season (*s*) we estimated disease burden and burden prevented by vaccination against B/Yamagata by multiplying our previous estimates for all influenza B lineages by the corresponding prevalence of B/Yamagata (*p*_*a,s*_). For example, if *H*_*a,s*_ is the estimated number of hospitalizations attributed to influenza B for age group *a* in season *s*, the number attributable to B/Yamagata was estimated as *H*_*a,s*_ × *p*_*a,s*_. Similarly, if *HV*_*a,s*_ is the estimated number of hospitalizations prevented by vaccination against influenza B, the number prevented by vaccination against B/Yamagata was estimated as *HV*_*a,s*_ × *p*_*a,s*_. To estimate burden that could have been caused by B/Yamagata in the absence of vaccination, we added the burden attributed to B/Yamagata with the estimated burden prevented by vaccination against B/Yamagata, for example, *p*_*a,s*_(*H*_*a,s*_ + *HV*_*a,s*_).

There was not a direct correspondence between the youngest age group of the B/Yamagata prevalence data and the influenza B burden estimates. We therefore assumed that B/Yamagata prevalence among children 0–4 years could approximate prevalence among children 6 months–4 years. Given the small sample size among age groups in some seasons (Table S1), we conducted an additional sensitivity analysis in which one population-wide value of B/Yamagata prevalence was applied across all ages. We also assumed that VE against influenza B was approximately equal to VE against B/Yamagata, so that estimates of burden prevented by vaccination against influenza B could be used to infer burden prevented by vaccination against B/Yamagata.

Finally, we constructed uncertainty distributions for the B/Yamagata burden estimates. First, we generated uncertainty around estimates of B/Yamagata prevalence using a Monte Carlo simulation approach described previously^9,10,11^. Briefly, we assumed prevalence for each age group and season followed a Binomially distributed random variable with probability of success equal to *p*_*a,s*_. We generated 10,000 independent samples from this distribution, 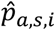, and multiplied these by the Monte Carlo samples previously generated for each influenza B burden estimate (for example, 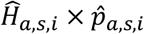, for *i* in 1, 2, … 10,000). We describe uncertainty as the 95^th^ percentiles of the resulting distributions. All in-text estimates are reported to three significant figures.

## Results

### B/Yamagata circulation from 2016/17–2019/20

The prevalence of B/Yamagata varied widely across seasons (Figure 1, Figure S1). B/Yamagata was most prevalent in 2016/17 and 2017/18, comprising approximately 16–22% of all influenza virus specimens with a known subtype or lineage (compared to 1% in 2018/19 and 2019/20). Among all influenza B virus specimens with known lineage (Table S1), there was an increase in B/Yamagata prevalence by age, and in every season the greatest proportions of B/Yamagata positive specimens were identified among adults 50–64 years and ≥65 years (Figure 1). Overall, the 2016/17 to 2019/20 seasons captured periods of high and low B/Yamagata circulation in addition to moderate to high epidemic severity, and thus represented a variety of plausible epidemiological scenarios.

**Figure 1.**
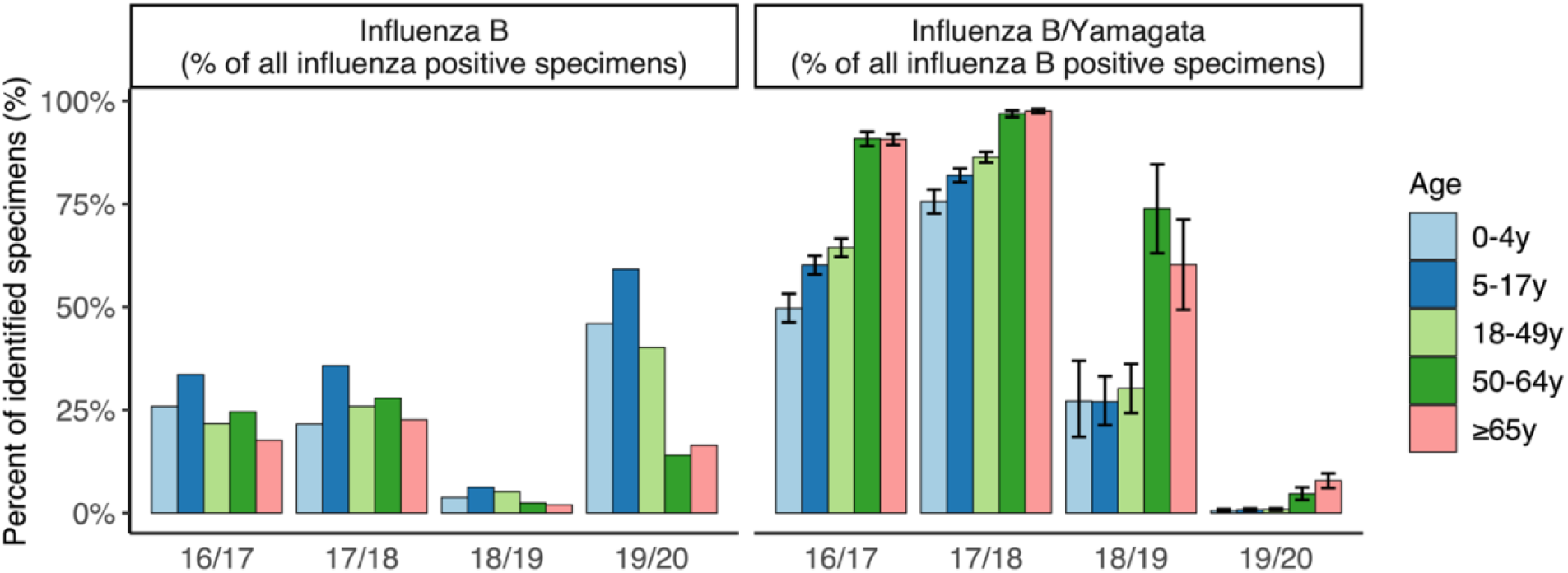
Prevalence of influenza B/Yamagata viruses. (Top) Percentage of all influenza A and B viruses with a known subtype or lineage that were identified as influenza B. (Bottom) Percentage of all influenza B viruses with a known lineage that were identified as influenza B/Yamagata. Error bars represent 95^th^ percentiles of the simulated uncertainty distributions.

### Estimates of B/Yamagata burden with and without vaccination

Across all seasons, we estimated that 22.9 million (95^th^ percentile uncertainty interval: 20.5–28.9 million) illnesses, 10.6 million (9.47–13.3 million) medically-attended illnesses, 340,000 (285,000–486,000) hospitalizations, and 25,100 (17,400–47,800) deaths would have occurred due to B/Yamagata in the absence of vaccination. Of these, vaccination prevented an estimated 4.15 million (3.51–5.45 million) illnesses, 2.01 million (1.68–2.65 million) medically-attended illnesses, 58,500 (40,900–91,300) hospitalizations, and 4,070 (2,300–8,270) deaths. Unsurprisingly, the greatest burden and prevented burden was estimated for the 2016/17 and 2017/18 seasons when B/Yamagata was most prevalent (Figure 2A–B). In particular, we estimated that a total of 217,000 (169,000–334,000) hospitalizations and 15,900 (9,010–36,200) deaths would have occurred during 2017/18 in the absence of vaccination.

**Figure 2.**
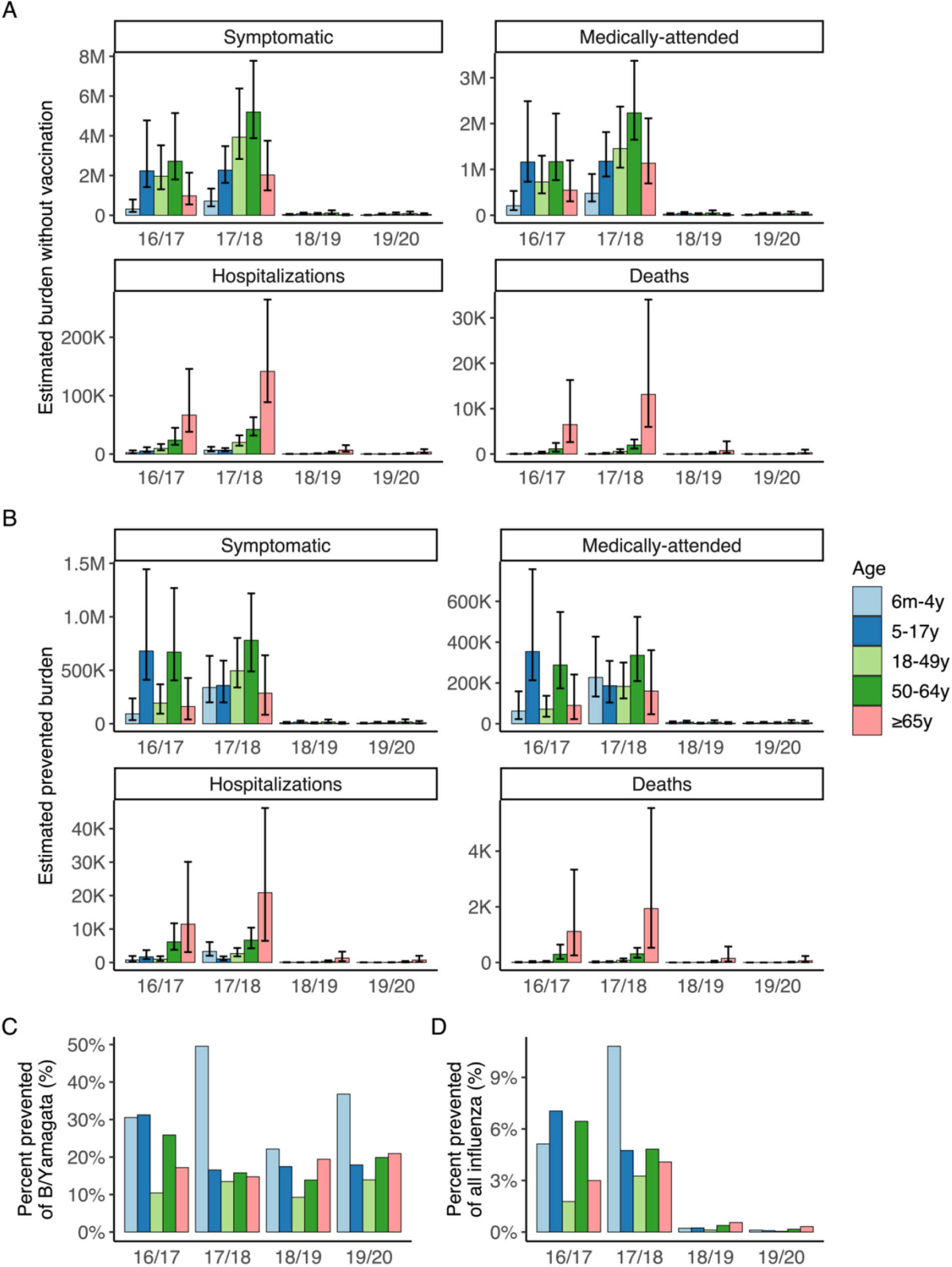
Estimated impact of vaccination against influenza B/Yamagata. (A) Estimates of disease burden in the absence of vaccination. (B) Estimates of disease burden prevented by vaccination. For (A) and (B), bars show point estimates and error bars show 95^th^ percentile uncertainty intervals. (C) Percentage of B/Yamagata hospitalizations prevented by vaccination. (D) Percentage of all influenza hospitalizations averted that were prevented by vaccination against B/Yamagata.

Among age groups, adults ≥65 years were estimated to experience the greatest burden of severe disease (hospitalizations and deaths) in the absence of vaccination against B/Yamagata, with up to 142,000 (88,800–264,000) hospitalizations and 13,100 (5,980–34,000) deaths occurring in a single season (2017/18). Results were similar when expressed as the rate of severe disease per 100,000 population (Figure S2). Vaccination also prevented the most severe disease among adults ≥65 years. For example, in 2017/18, vaccination prevented 20,900 (6,460–46,200) hospitalizations and 1,940 (530– 5,550) deaths due to B/Yamagata. Conversely, vaccination generally prevented the greatest percentage of B/Yamagata hospitalizations among children 6 months–4 years (22–50% across seasons; Figure 2C). This corresponded to up to 11% of all influenza-associated hospitalizations among young children, depending on the season (Figure 2D). Results were similar when we used one B/Yamagata prevalence estimate for all age groups, rather than age-specific estimates (Figure S3). Overall, our results suggest older adults may be at greatest risk of severe disease if B/Yamagata were to recirculate in the absence of vaccine-mediated protection.

## Discussion

We estimated the impact of vaccination against B/Yamagata-associated outcomes by combining data on B/Yamagata prevalence from 2016–2020 with estimates of influenza B-associated illnesses, hospitalizations and deaths prevented by vaccination during the same period. Our estimates of burden prevented by vaccination, and burden that would have occurred without vaccination, were greatest in 2016/17 and 2017/18 due to high prevalence of B/Yamagata during those seasons. For all seasons, vaccination prevented the most severe disease among adults ≥65 years, who experienced the highest prevalence of B/Yamagata and are at increased risk of severe complications following influenza virus infection. In contrast, the greatest percent of severe disease was prevented among young children. Overall, our results highlight the importance of continued B/Yamagata surveillance, and readiness to respond to possible re-emergence, given the potential for substantial morbidity and mortality without vaccination, particularly among groups at increased risk of severe influenza-associated complications.

Our estimates of the impact of vaccination in mitigating the burden of influenza B/Yamagata can help inform assessments of the potential impacts if B/Yamagata were to return to circulation in the absence of vaccination-mediated protection. In seasons with high circulation of B/Yamagata, we estimated up to 217,000 hospitalizations and 15,900 deaths would have occurred in the absence of vaccination, including up to 142,000 hospitalizations and 13,100 deaths among adults ≥65 years. Since our estimates reflect disease burden in the presence of some level of population immunity to B/Yamagata, which may have declined since 2020 and/or may decline with its continued absence from circulation and removal from the vaccine, morbidity and mortality could be higher than we have reported. However, any realized burden will depend on many factors, including the prevalence of B/Yamagata relative to other influenza viruses, the strength of cross-protection induced by vaccination against B/Victoria, and levels and durations of pre-existing immunity to B/Yamagata conferred through prior infection or vaccination.

There are several limitations that should be considered in interpreting these results. First, we assumed VE against all influenza B viruses was reflective of VE against influenza B/Yamagata. This was likely true in 2016/17 and 2017/18 when B/Yamagata was the dominant influenza B lineage. Conversely, the prevalence of B/Yamagata was so low in 2018/19 and 2019/20 that a small change in VE is unlikely to have a substantial impact on our absolute estimates of burden prevented by vaccination. Second, information on influenza vaccination was self-reported for adults and parent-reported for children and may be subject to recall bias. However, self-reported seasonal influenza vaccination status for adults has been shown to have relatively high agreement with vaccination status ascertained from medical records^12^. Finally, we assumed all vaccinees received a formulation containing B/Yamagata. However, trivalent vaccines in 2016/17–2019/20 did not include B/Yamagata and were still in use in the United States, albeit at declining levels relative to quadrivalent vaccines (for example, in 2017/18, 97% of Flu VE Network participants aged <65 years with known vaccine type received a quadrivalent inactivated vaccine)^9,13–15^. Data on trivalent vaccination coverage were not available for all age groups and seasons to directly account for this in our framework, and our estimates of burden prevented by vaccination, and thus our estimates of burden without vaccination, may be overestimated if there was less vaccine-mediated cross-protection against B/Yamagata than we have assumed. However, our methods are also conservative in that they do not account for indirect effects of vaccination in reducing infection risks through herd immunity.

We have found that vaccination against B/Yamagata prevented substantial morbidity and mortality during seasons in which the virus circulated at high levels, particularly for adults ≥65 years. Continued surveillance for B/Yamagata recirculation will be important given its potential to cause substantial morbidity and mortality in the absence of vaccination.

## Funding

This research did not receive any specific grant from funding agencies in the public, commercial, or not-for-profit sectors.

## Conflicts of Interest

The authors have no competing interests to declare.

## Author Contributions

All authors attest they meet the ICMJE criteria for authorship.

## Data availability statement

All data were the result of secondary analyses. The majority are publicly available and cited within in the text. A small portion are not publicly available due to privacy restrictions but are available upon reasonable request to the authors.

## Notes

### Competing Interest Statement

The authors have declared no competing interest.

### Author Declarations

All data were the result of secondary analyses. The Centers for Disease Control and Prevention determined that the study met the requirement for public health surveillance, and institutional review board (IRB) approval and patient consent were not required.

