## Supplementary Information for "Estimating historical impacts of vaccination against influenza B/Yamagata in the United States to inform possible risks of re-emergence in the absence of vaccination"

**Table S1** – Total number of influenza B positive specimens with known lineage obtained from FluView.

| Age | 2016/17 | 2017/18 | 2018/19 | 2019/20 |
|---|---|---|---|---|
| 0-4y | 789 | 824 | 92 | 2361 |
| 5-17y | 1846 | 1972 | 211 | 4642 |
| 18-49y | 1794 | 2696 | 235 | 4923 |
| 50-64y | 1072 | 1960 | 65 | 689 |
| ≥65y | 1828 | 3429 | 73 | 905 |

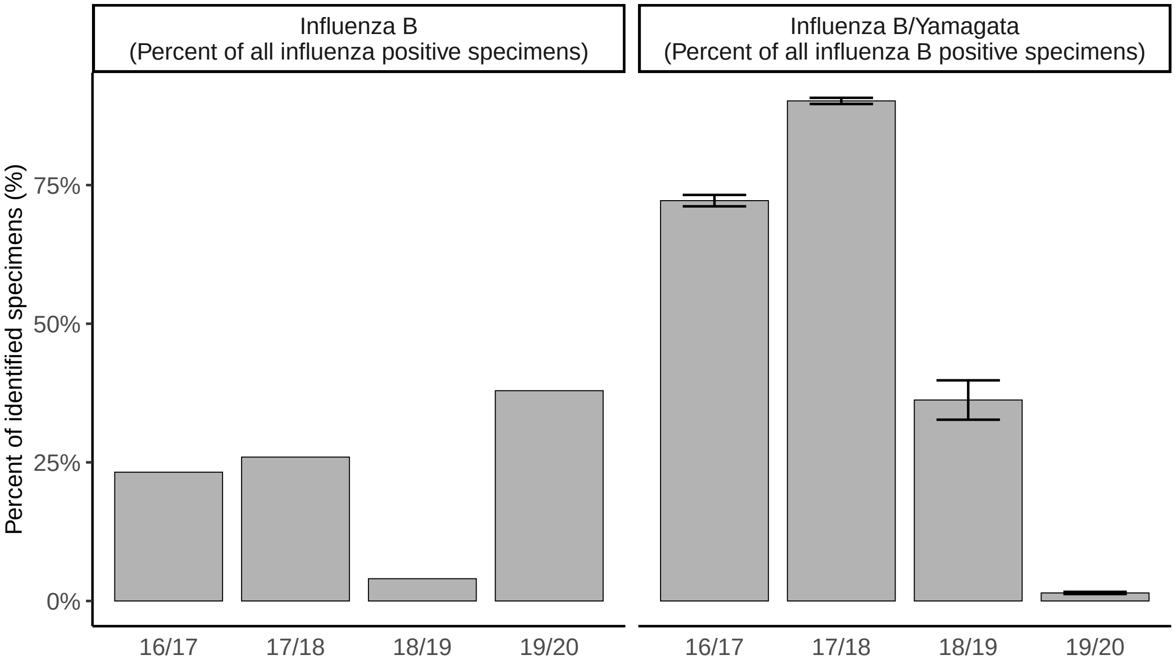


**Figure S1** – **Prevalence of influenza B/Yamagata viruses across all ages.** (Left) Percentage of all influenza A and B viruses with a known subtype or lineage that were identified as influenza B. (Right) Percentage of all influenza B viruses with a known lineage that were identified as influenza B/Yamagata. Error bars represent 95th percentiles of the simulated uncertainty distributions.

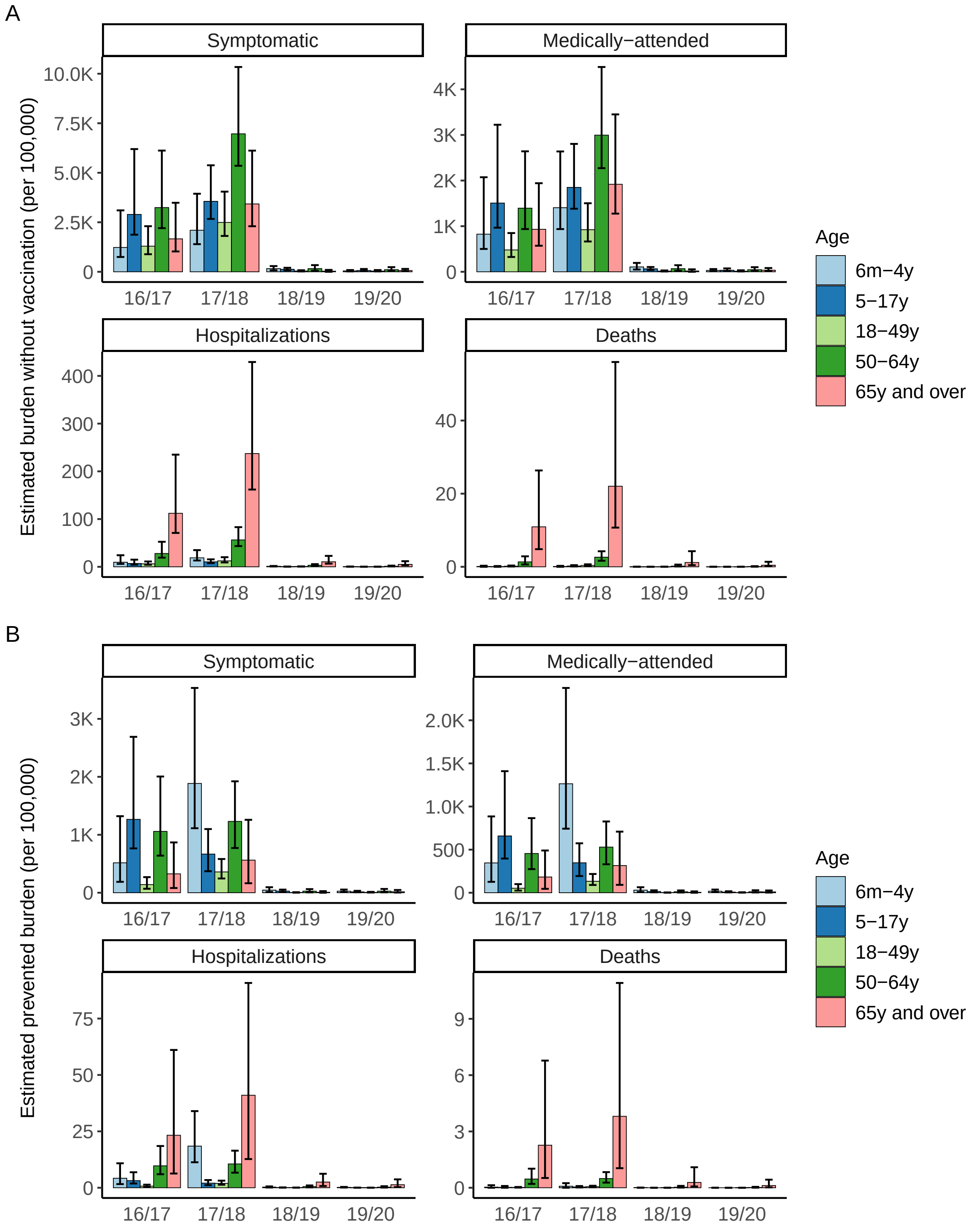


**Figure S2 – Estimated impact of vaccination against influenza B/Yamagata per 100,000 population.** (A) Estimates of disease burden in the absence of vaccination. (B) Estimates of disease burden prevented by vaccination. For (A) and (B), bars show point estimates and error bars show 95th percentile uncertainty intervals.

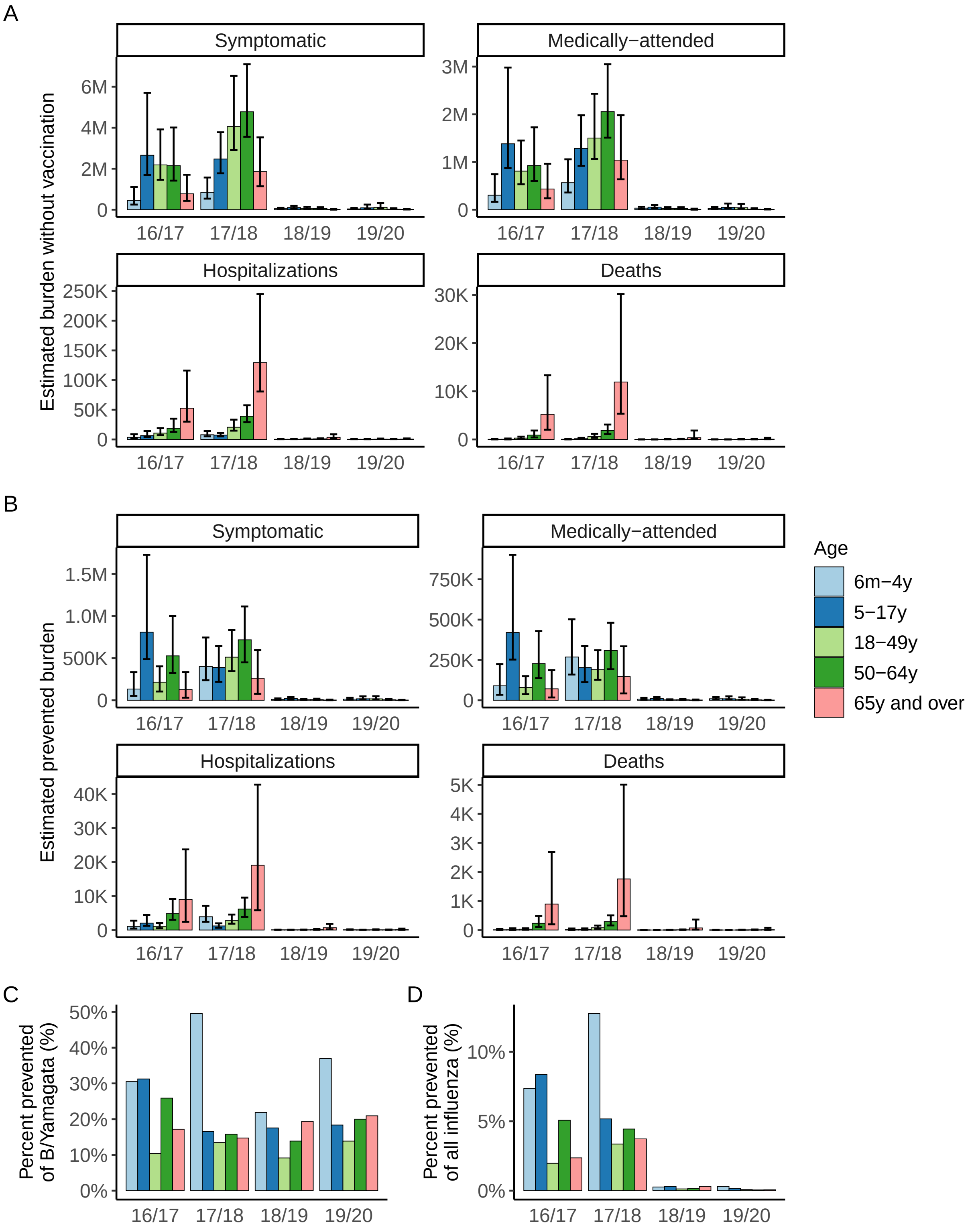


**Figure S3 – Estimated impact of vaccination against influenza B/Yamagata when one prevalence estimate is used for all age groups.** (A) Estimates of disease burden in the absence of vaccination. (B) Estimates of disease burden prevented by vaccination. For (A) and (B), bars show point estimates and error bars show 95$^{th}$ percentile uncertainty intervals. (C) Percentage of B/Yamagata hospitalizations averted by vaccination. (D) Percentage of all influenza hospitalizations averted that were averted by vaccination against B/Yamagata.
